# OASIS+: leveraging machine learning to improve the prognostic accuracy of OASIS severity score for predicting in-hospital mortality

**DOI:** 10.1101/2020.12.28.20248946

**Authors:** Yasser EL-Manzalawy, Mostafa Abbas, Ian Hoaglund, Alvaro Ulloa Cerna, Thomas B. Morland, Christopher M. Haggerty, Eric S. Hall, Brandon K. Fornwalt

## Abstract

Severity scores assess the acuity of critical illness by penalizing for the deviation of physiologic measurements from normal and aggregating these penalties (also called “weights” or “subscores”) into a final score (or probability) for quantifying the severity of critical illness (or the likelihood of in-hospital mortality). Although these simple additive models are human readable and interpretable, their predictive performance needs to be further improved. To address this need, we argue for replacing these simple additive models with models based on state-of-the-art non-linear supervised learning algorithms (e.g., Random Forest (RF) and eXtreme Gradient Boosting (XGB)). Specifically, we present OASIS+, a variant of the Oxford Acute Severity of Illness Score (OASIS) in which an ensemble of 200 decision trees is used to predict in-hospital mortality based on the 10 same clinical variables in OASIS. Using a test set of 9566 admissions extracted from MIMIC-III database, we show that the performance of OASIS can be substantially improved from AUC score of 0.77 to 0.83 using OASIS+. Moreover, we show that OASIS+ has superior performance compared to eight other commonly used severity scoring methods. Our results underscore the potential of improving existing severity scores by using more sophisticated machine learning algorithms (e.g., ensemble of non-linear decision tress) not just via including additional physiologic measurements.

## Background

In the past three decades, several severity scores have been developed with the primary objective of predicting in-hospital mortality from clinical and/or biological measurements, often collected within the first 24 hours of admission to the intensive care unit (ICU) admission [1, 2]. Point-based severity scores compute the severity of an illness by modeling the deviations of a set of clinical and/or biological variables from their normal physiologic values [2]. More specifically, point-based severity scores rely on linear or non-linear transformations of the measurements into subscores from which a final score is computed as a linear sum. Based on this approach, several severity scoring methods have been proposed including Acute Physiology and Chronic Health Evaluation (APACHE) [3-6], Simplified Acute Physiology Score (SAPS) [7-10], Logistic Organ Dysfunction Score (LODS) [11], Systemic Inflammatory Response Syndrome (SIRS) [12], Sequential Organ Failure Assessment (SOFA) [13], and Oxford Acute Severity of Illness Score (OASIS) [14]. These severity scoring methods have been widely used in many research and clinical applications including predicting mortality, length of stay (LoS), stratifying patients for clinical trials, and evaluation of ICU quality of care [15].

Early prediction of mortality in ICU patients can improve health outcomes and is essential for timely interventions by ICU clinicians [16]. Regardless of the availability of many severity scoring methods, the prognostic performance of these models in predicting in-hospital mortality remains far from satisfactory [17, 18]. Since these severity scoring methods can be viewed as logistic regression models, one promising direction for boosting their predictive performance is to replace the simple additive model with more sophisticated supervised machine learning algorithms such as Random Forest (RF) [19] or eXtreme Gradient Boosting (XGB) [20]. Unfortunately, the expected improvement in performance might introduce a tradeoff in model interpretability resulting from the increased model complexity which might also hamper clinicians’ ability to associate explanations with the predictions made by the model.

Against this background, our preliminary objective is to accentuate the promise and demonstrate the feasibility of developing a new generation of severity scoring methods based on state-of-the-art non-linear supervised learning algorithms. Specifically, we present OASIS+, a novel method for predicting in-hospital mortality using an ensemble of non-linear decision trees trained using the same 10 clinical variables used to calculate the OASIS score. Using a test set of ICU stays lasting at least for 24 hours that represent 9566 distinct adult patients, we show that OASIS+ outperforms nine previously developed severity scoring methods (including OASIS) in predicting in-hospital mortality.

## Methods

### Ethics statement

The retrospective cohort training and test datasets were extracted from the Medical Information Mart for Intensive Care III (MIMIC-III) database (version 1.4) [21]. MIMIC-III is a publicly available database of 46,476 ICU patients hospitalized in Beth Israel Deaconess Medical Center (BIDMC) and the public access to the de-identified database has been approved by the BIDMC and MIT institutional review boards [21]. All data processing and analyses presented in this study have been conducted in accordance with MIMIC-III guidelines and regulations.

### Study population

MIMIC-III contains clinical data for 49,785 hospital admissions associated with 38,597 adult patients admitted to ICU between 2001 and 2012. The following criteria were used to exclude patients from our cohort dataset: i) age less than 18 years or greater than 90 years; ii) ICU stays with duration less than 24 hours. For patients with multiple ICU stays, we included the first ICU stay with LoS greater than or equal to 24 hours. The primary outcome for our analysis was in-hospital mortality. Our final dataset included 31,884 distinct patients and ICU stays. We randomly partitioned the data into 70% and 30% for training and testing, respectively. Missing variables were imputed as normal [22] such that their corresponding missing subscores were ZEROs.

### Data extraction

We downloaded MIMIC-III files from https://mimic.physionet.org/. Then, we followed the instructions from the MIMIC code repository [23] to build a local PostgreSQL [24] database and adapted MIMIC-III code to extract our dataset and compute patient comorbidities and first day severity scores. For supporting reproducibility of our work, the PostgreSQL query script is provided in the supplementary material.

### Point-based severity scores

Since their presentation in the 1980s, point-based severity scores (e.g., APACHE-II, SAPS-III, OASIS) have been commonly used in the ICU settings for assessing disease severity in critically ill patients and predicting poor health outcomes such as in-hospital or 30-day mortality [15]. Many severity scores are calculated using clinical variables (e.g., temperature and heart rate) and biological variables (e.g., white blood cell count) collected from the first day in the ICU (see Table S1). Other severity scores such as (SOFA) [13] can be computed repetitively every day or every time new measurements are presented.

A common characteristic among point-based severity scores is that they are based on logistic regression models and, therefore, can be viewed as multivariate additive linear models for predicting the severity of critical illness. Such models can be easily computed and interpreted by a physician. Briefly, each variable is transformed into a subscore such that a subscore of ZERO indicates that the measured variable is within its normal range and higher subscores penalize for observed variables outside their normal range (or value). These subscores are determined using a consensus opinion or data-driven approaches [1]. An overall severity score is then computed as the sum of all subscores. The higher the overall score, the greater the disease burden and the higher likelihood of poor health outcomes. For example, let’s consider the OASIS severity score [14]. This score is computed using 10 clinical variables from first day in the ICU. Using Table 1, each clinical variable is transformed into a corresponding subscore and the OASIS score is the sum of these 10 subscores. An OASIS probability of mortality is also computed using 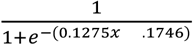 where *x* is the OASIS score.

**Table 1:**
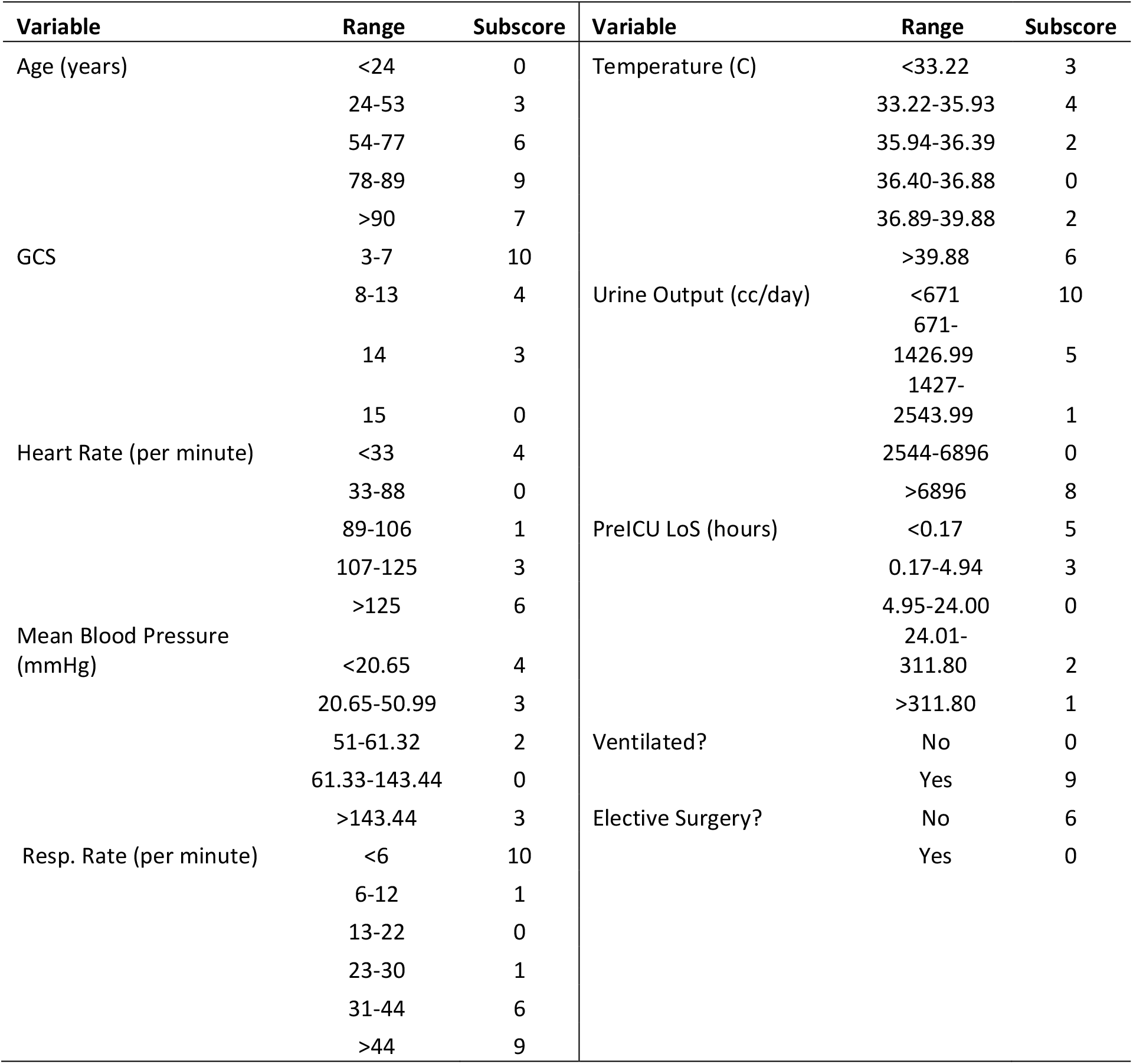
Mapping OASIS clinical variables into subscores.

In the present study, we assessed the predictive performance of nine previously developed severity scores (summarized in Table S1) for predicting in-hospital mortality using data collected during the first ICU day.

### Machine learning models

We experimented with three widely used supervised machine learning algorithms: i) Random Forest [19] with 200 trees (RF200); ii) eXtreme Gradient Boosting [20] with 200 weak tree learners (XGB200); iii) Logistic Regression (LR) [25] with L2 regularization. These algorithms are implemented in the Scikit-learn machine learning library [26]. The LR model is a linear and human interpretable model while RF200 and XGB200 models are ensembles of 200 non-linear decision trees. To get insights into how these ensemble models work, we used feature importance scores to quantify the contribution of each feature to the predictions made by both RF200 and XGB200.

### Statistical analysis and performance evaluation

Categorical variables are reported as percentages and the chi-square test was used to assess whether two (or more) proportions are different from each other. Continuous variables are summarized as medians and interquartile ranges (IQR) and the non-parametric Mann-Whitney test was used to determine the differences in the distribution of a variable (e.g., Age) in survival and non-survival groups. All statistical analyses were performed using R version 3.6.2 [27] and a p-value less than 0.05 was considered significant.

The predictive performance of the machine learning models was assessed using five commonly used predictive performance metrics [28]: Accuracy (ACC), Sensitivity (Sn); Specificity (Sp); Matthews correlation coefficient (MCC); and Area Under the receiver operator Curve (AUC) [29]. The prognostic performance of different models is assessed using calibration curves [30] and the root-mean-square error (RMSE) is used to quantify calibration errors [31].

## Results

### Characteristics of train and test sets

Basic summary statistics of the training and test sets are provided in Tables 2 and S2, respectively. Both tables show that the following variables are significantly associated with in-hospital mortality in ICU patients: increased age, increased length of stay, increased number of pre-existing conditions (i.e., comorbidities), and increased severity of the critical illness (using any of the nine pre-existing severity scores considered in our analysis). Although this exploratory analysis suggests that any randomly selected patient from the non-survival group is likely to have severity scores higher than those for a randomly selected patient from the survival group, it is of particular interest to assess how well these severity scores can discriminate between patients within each of the two groups.

**Table 2:**
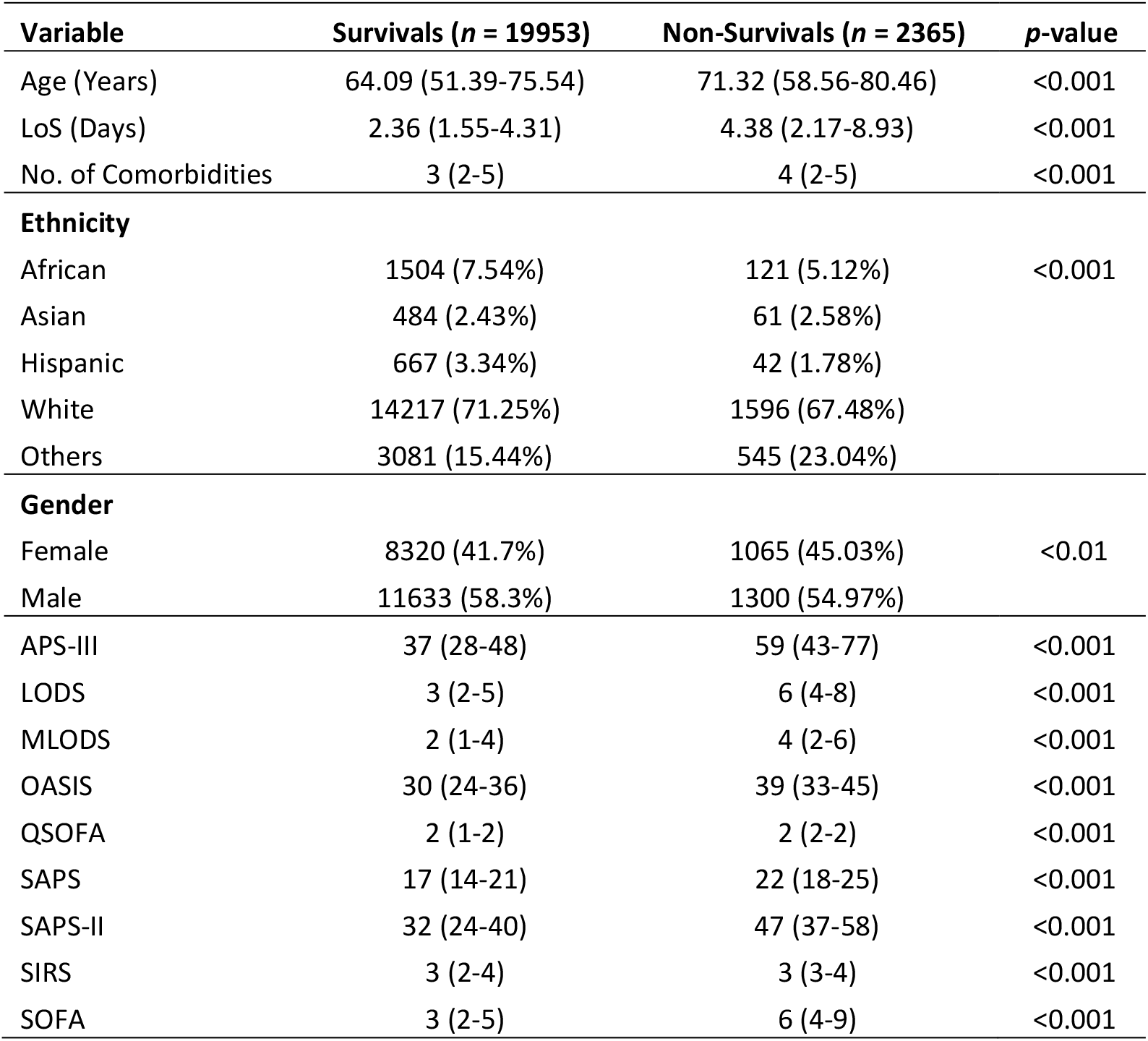
Summary statistics of MIMIC-III training data.

| Variable | Survivals (*n* = 19953) | Non-Survivals (*n* = 2365) | *p*-value |
| --- | --- | --- | --- |
| Age (Years) | 64.09 (51.39-75.54) | 71.32 (58.56-80.46) | <0.001 |
| LoS (Days) | 2.36 (1.55-4.31) | 4.38 (2.17-8.93) | <0.001 |
| No. of Comorbidities | 3 (2-5) | 4 (2-5) | <0.001 |
| **Ethnicity** |  |  |  |
| African | 1504 (7.54%) | 121 (5.12%) | <0.001 |
| Asian | 484 (2.43%) | 61 (2.58%) |  |
| Hispanic | 667 (3.34%) | 42 (1.78%) |  |
| White | 14217 (71.25%) | 1596 (67.48%) |  |
| Others | 3081 (15.44%) | 545 (23.04%) |  |
| **Gender** |  |  |  |
| Female | 8320 (41.7%) | 1065 (45.03%) | <0.01 |
| Male | 11633 (58.3%) | 1300 (54.97%) |  |
| APS-III | 37 (28-48) | 59 (43-77) | <0.001 |
| LODS | 3 (2-5) | 6 (4-8) | <0.001 |
| MLODS | 2 (1-4) | 4 (2-6) | <0.001 |
| OASIS | 30 (24-36) | 39 (33-45) | <0.001 |
| QSOFA | 2 (1-2) | 2 (2-2) | <0.001 |
| SAPS | 17 (14-21) | 22 (18-25) | <0.001 |
| SAPS-II | 32 (24-40) | 47 (37-58) | <0.001 |
| SIRS | 3 (2-4) | 3 (3-4) | <0.001 |
| SOFA | 3 (2-5) | 6 (4-9) | <0.001 |

Table 3 characterizes the 10 clinical variables used for computing OASIS severity scores in the training and test sets. For both datasets, all clinical variables (except pre-ICU LoS) were found to be drawn from significantly different distributions for survival and non-survival groups. It should be noted that the non-parametric Mann-Whitney test is a rank sum test that ranks all of the observations from each group and then sums the ranks from one of the groups which is compared with the expected rank sum. Therefore, it is possible for the two groups to have significantly different rank sums while their medians are equal as we noted for the Glasgow Coma Score (GCS) and Pre-ICU LoS variables.

**Table 3:** Summary statistics of OASIS variables in train and test sets.

| Variable | Train (*n* = 22318) |  |  | Test (*n* = 9566) |  |  |
| --- | --- | --- | --- | --- | --- | --- |
|  | Survivals (*n* = 19953) | Non-Survivals (*n* = 2365) | *p*-value | Survivals (*n* = 8560) | Non-Survivals (*n* = 1006) | *p*-value |
| Age | 64 (51-75) | 71 (58-80) | <0.001 | 63 (51-75) | 71.5 (60-80) | <0.001 |
| GCS | 15 (14-15) | 15 (13-15) | <0.001 | 15 (14-15) | 15 (12-15) | <0.001 |
| Heart Rate | 101 (90-115) | 111 (95-128) | <0.001 | 101 (90-115) | 110 (94-126) | <0.001 |
| Mean Blood Pressure | 59.67 (52.67-86) | 55 (47-82) | <0.001 | 59 (53-86) | 55 (47-81.21) | <0.001 |
| Respiratory Rate | 26 (18.5-30) | 28 (24-33) | <0.001 | 26 (18-30) | 28 (24-33) | <0.001 |
| Temperature | 36.17 (35.7-37.5) | 36 (35.44-37.5) | <0.001 | 36.17 (35.72-37.5) | 36.06 (35.5-37.71) | <0.001 |
| Urine Output | 1895 (1245-2770) | 1189 (645-2050) | <0.001 | 1900 (1235-2785) | 1167 (640.5-1972.5) | <0.001 |
| Pre-ICU-LoS | 0 (0-18) | 0 (0-23) | <0.001 | 0 (0-17) | 0 (0-27) | 0.12 |
| Ventilated?: Yes | 9723 (48.73%) | 1570 (66.38%) | <0.001 | 4186 (48.9%) | 676 (67.2%) | <0.001 |
| Elective Surgery?: Yes | 3357 (16.82%) | 76 (3.21%) | <0.001 | 1367 (15.97%) | 38 (3.78%) | <0.001 |

Table S3 summarizes the baseline characteristics of Elixhauser comorbidities [32] for the survival and non-survival groups in the training and test datasets. Out of the 30 Elixhauser comorbidities, 18 comorbidities in the training set and 16 comorbidities in the test set have significantly different proportions of survivals and non-survivals. The top three most frequent comorbidities are: Hypertension (more frequent in survivals), Fluid and Electrolyte Disorders (more frequent in non-survivals), and Cardiac Arrhythmias (more frequent in non-survivals).

### Assessment of nine severity scores on predicting in-hospital mortality

We evaluated the performance of nine previously developed severity scores (Table S1) on predicting in-hospital mortality using the MIMIC-III test set. Table 4 presents the performance of these nine severity scores using five widely used metrics. Since every scoring method has different scale, we normalized each score in the range [0,1] and we used the MIMIC-III training set to estimate the optimal threshold for transforming the scores into binary labels (i.e., survivals vs. non-survivals). The optimal threshold was computed by maximizing the Youden’s J statistic [33]. Fig. 1 shows the Receiver Operating Curve (ROC) curves and corresponding AUC scores for each of the nine severity score methods. We found that models with the best performance (i.e., *AUC* ∈ [0.77 − 0.8]) were based on OASIS, APS-III, and SAPS-II. SOFA, MLODS, SAPS, and LODS demonstrated moderate performance (i.e., *AUC* ∈ [0.72 − 0.75]), and SIRS and QSOFA performed poorly with AUC scores of 0.61 and 0.59, respectively. Not only did the SAPS-II model have the highest AUC, but its ROC curve demonstrated superior performance (in terms of threshold-dependent metrics) at all possible thresholds compared to the ROC curves for the other eight severity models.

**Table 4:**
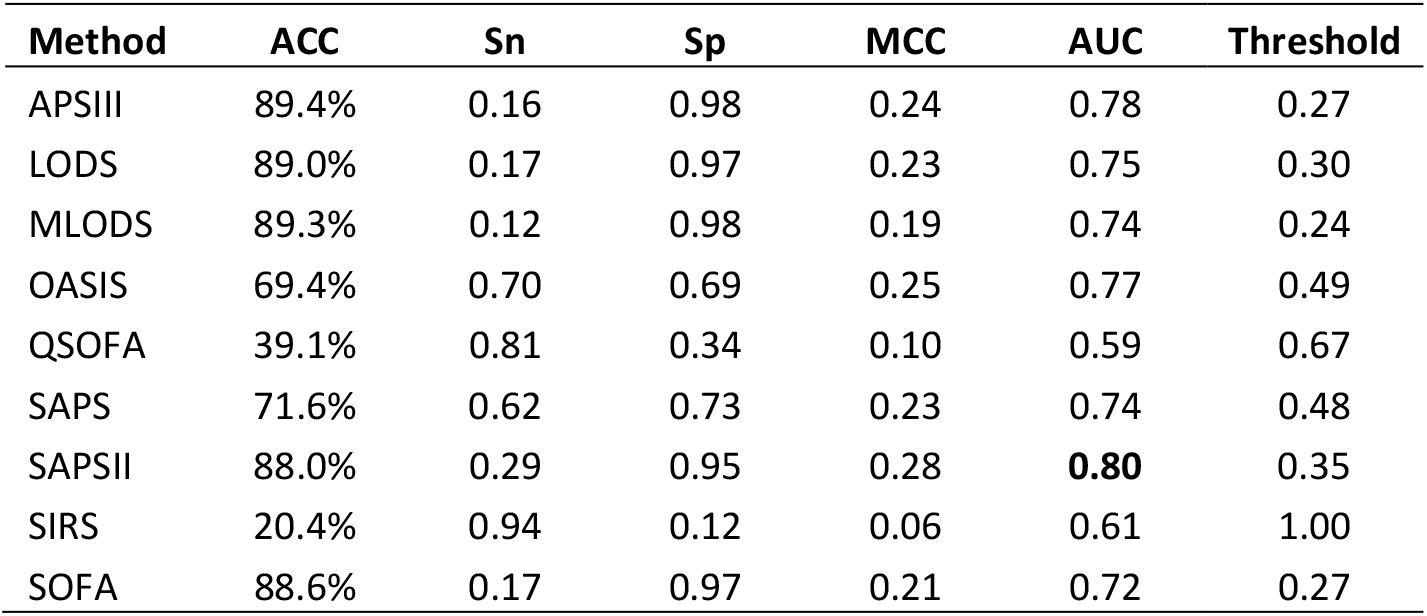
Performance comparisons of nine severity score models for predicting in-hospital mortality estimated using MIMIC-III test set.

**Fig. 1.**
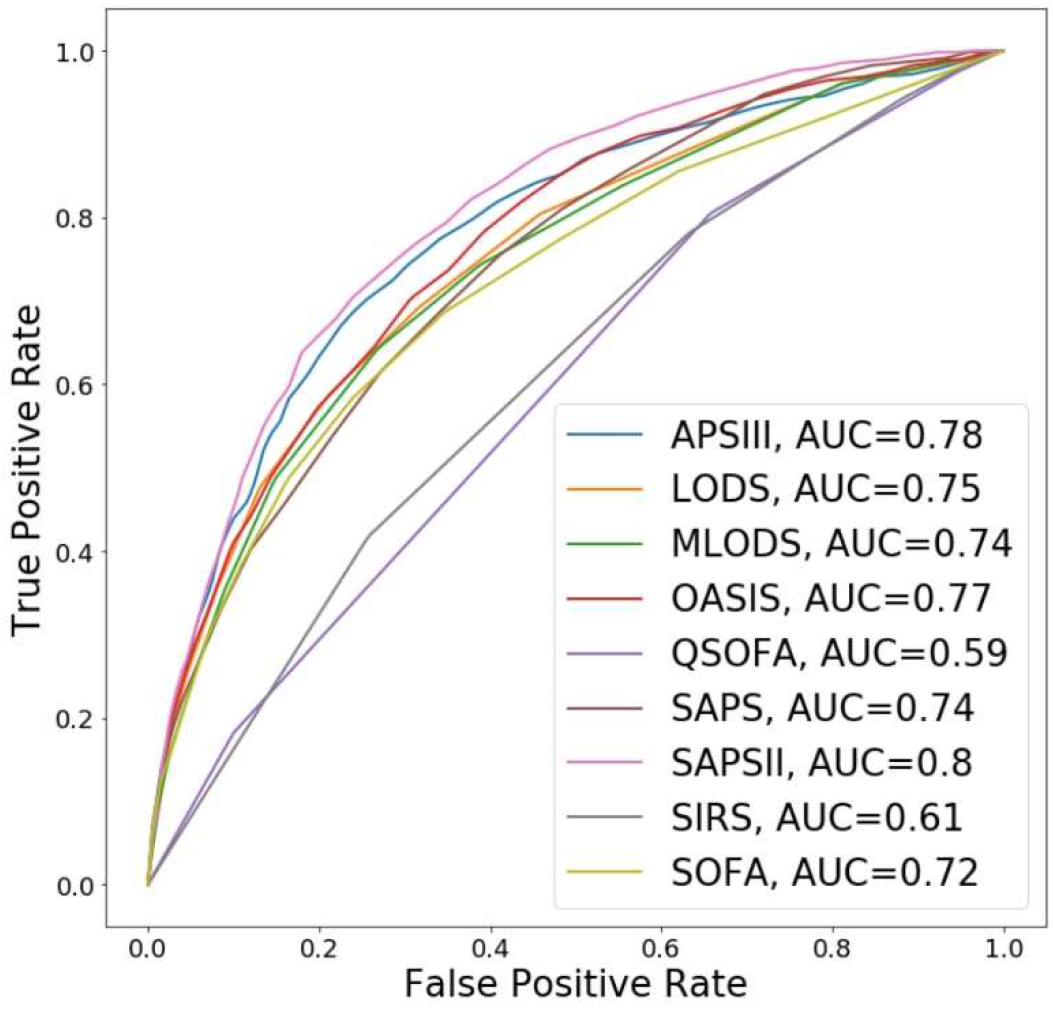
Performance (in terms of ROC curves and associated AUC scores) of nine severity scores estimated using MIMIC-III test set for predicted in-hospital mortality.

Of the top three severity scores, SAPS-II and APS-III scores are computed using 14 and 20 variables, respectively (See Table S1), and OASIS score is computed using 10 variables (See Table 1). Therefore, among these three scores, OASIS score: uses the smallest number of variables; uses no biological variables; and has the lowest performance on predicting in-hospital mortality. In what follows, we present a novel variant of OASIS score, OASIS+, that leverages non-linear machine learning supervised algorithms for outperforming the SAPS-II and APS-III models on predicting in-hospital mortality.

### OASIS+ outperforms all nine severity scores on predicting in-hospital mortality

We considered two approaches for developing supervised learning classifiers using the 10 clinical variables used for computing OASIS scores (Table 1). In the first approach, we used the 10 subscores and OASIS probability as input features. In the second approach, we used the 10 clinical variables (without transforming them into subscores) as input features. In both cases, we evaluated one linear supervised learning algorithm, LR, and two algorithms for building ensembles of 200 non-linear decision trees, RF200 and XGB200. The predictive performance of these six models is summarized in Fig. 2 and Table 5. Using OASIS subscores, the best performing model, XGB200, has an AUC score of 0.81 while models based on LR and RF200 slightly outperform OASIS score. Using OASIS variables, RF200 and XGB200 have AUC scores of 0.82 and 0.83, respectively. The results show that the two ensemble learning algorithms achieve better performance when trained using the measured values of the clinical variables as opposed to their non-linear transformation derived from OASIS benchmark dataset [14]. This observation suggests that OASIS non-linear transformations in Table 1 are more likely to be data-specific and might not generalize well to other patient populations. The main interesting observation is that a 0.06 improvement in AUC score is obtained by replacing the OASIS linear additive scoring function with the non-linear XGB model trained using non-transformed clinical variables (hereafter called OASIS+ model).

**Table 5:** **Performance comparisons of different machine learning models for predicting in-hospital mortality estimated using MIMIC-III test set**

| **Features** | **Method** | **ACC** | **Sn** | **Sp** | **MCC** | **AUC** | **Threshold** |
| --- | --- | --- | --- | --- | --- | --- | --- |
| OASIS subscores | RF200 | 77.5% | 0.54 | 0.80 | 0.25 | 0.76 | 0.16 |
|  | LR | 73.9% | 0.66 | 0.75 | 0.28 | 0.78 | 0.12 |
|  | XGB200 | 70.9% | 0.79 | 0.70 | 0.31 | 0.81 | 0.10 |
| OASIS variables | RF200 | 88.0% | 0.34 | 0.94 | 0.31 | 0.82 | 0.33 |
|  | LR | 70.0% | 0.69 | 0.70 | 0.25 | 0.77 | 0.10 |
|  | XGB200 | 72.8% | 0.78 | 0.72 | 0.33 | 0.83 | 0.10 |

**Fig. 2.**
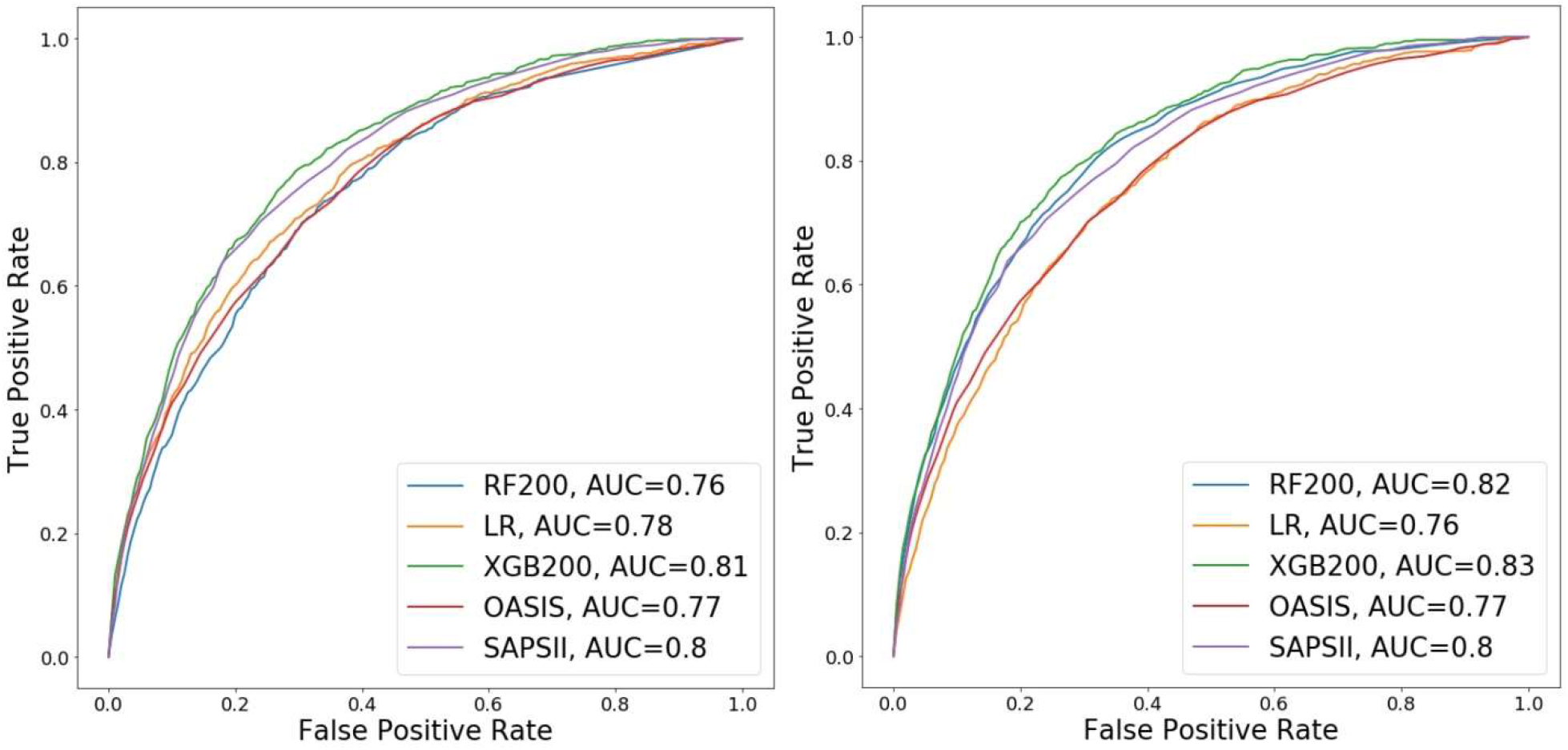
Performance (in terms of ROC curves and associated AUC scores) of three machine learning models for predicting in-hospital mortality trained using oasis score and subscores (left) and oasis variables (right).

Since OASIS+ is a prognostic model for predicting the risk of in-hospital mortality, it should be evaluated in terms of discrimination (e.g., using AUC and threshold-dependent metrics) as well as calibration [31, 34]. Fig. 6 shows the calibration curves for SAPS-II, OASIS, and the three machine learning models trained using OASIS variables. We found that OASIS and SAPS-II have higher estimated calibration errors when compared with our three machine learning models. The lowest estimated calibration error is observed for XGB200 (OASIS+) and RF200 models.

Fig. 3 shows the contribution of each variable in the predictions made by OASIS+ model also called OASIS+ feature importance scores. Comparing OASIS+ feature importance scores (Fig. 3) with OASIS non-linear transformations (Table 1) reveals several discrepancies between relative variable importance of the two models. For example, in OASIS+, the three variables with the highest feature importance scores are: elective surgery, ventilation use, and urine output. On the other hand, the three OASIS variables with the highest weights (i.e., OASIS subscore of 10) are: GCS in the range 3-7, respiratory rate less than 6 per minute, and urine output less than 671 cc/day. Unlike OASIS+, each variable in OASIS can take different weights with a ZERO weight corresponding to normal measurements. Thus, for a given case, respiratory rate might have the highest contribution (subscore of 10) to the computed OASIS score while in another case respiratory rate might be in the normal range (13-32) and have a ZERO contribution to the OASIS score.

**Fig. 3.**
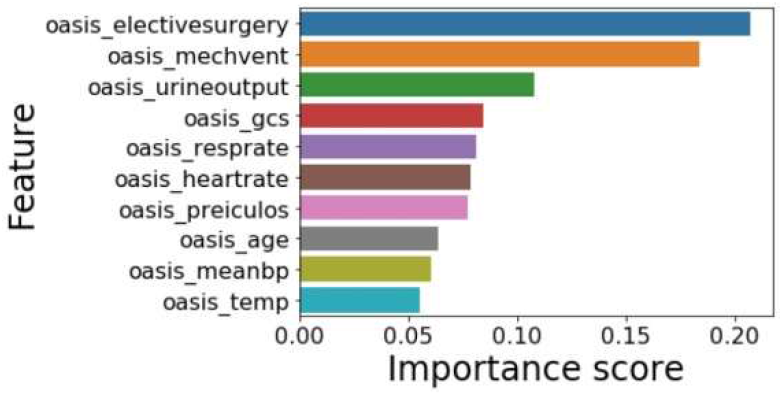
Features importance scores of the OASIS+ model.

Fig. 4 shows the violin plots, estimated using the test set, for normalized OASIS scores, OASIS probabilities, and OASIS+ predicted probabilities in survivals and non-survivals groups. A violin plot shows both a box plot and a rotated kernel density plot. We noted that OASIS scores follow a normal distribution. This acknowledges the same finding reported in [14] on a different patient population. In the three cases, the median score or probability in the non-survival group is higher than the corresponding median in the survival group. However, the largest and most significant (p-value equals 2.4e-250) difference between the two medians is observed for OASIS+ probabilities.

**Fig. 4.**
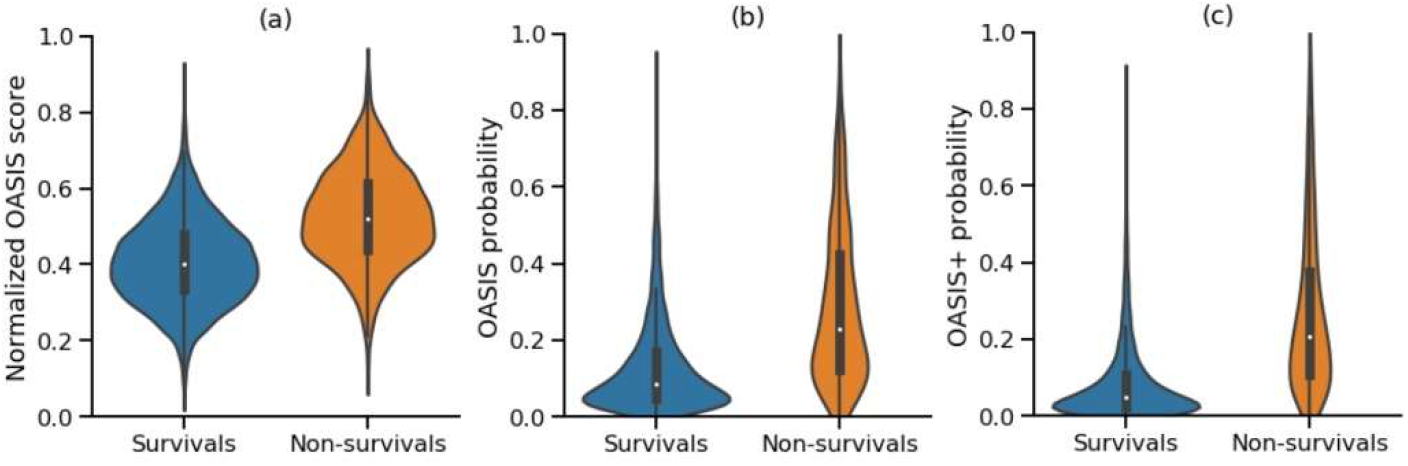
Violin plots, for (a) normalized OASIS scores (b) OASIS probabilities (c) OASIS+ probabilities in survivals and non-survivals groups, computed using the MIMIC-III test set.

Finally, Table 5 compares the performance of the six predictive models, considered in this experiment, using AUC and four threshold-dependent metrics. For all the models, we noticed that the optimal threshold, estimated using the training data only, for transforming the model predicted probability into a binary label is lower than 0.5. This is an artifact of the uneven ratio of non-survivals to survivals since the number of survival cases is almost 10 times the number of non-survivals. Fig. 5 shows the tradeoff between sensitivity and specificity in OASIS+ for different choices of the probability threshold. At a threshold equals 0.1, the sensitivity and specificity of the model are 0.78 and 0.72, respectively. The highest MCC of 0.36 is reached at a threshold of 0.13 which yields 0.70 and 0.80 sensitivity and specificity, respectively. The complete set of OASIS+ results at different thresholds is provided in Table S4.

**Fig. 5.**
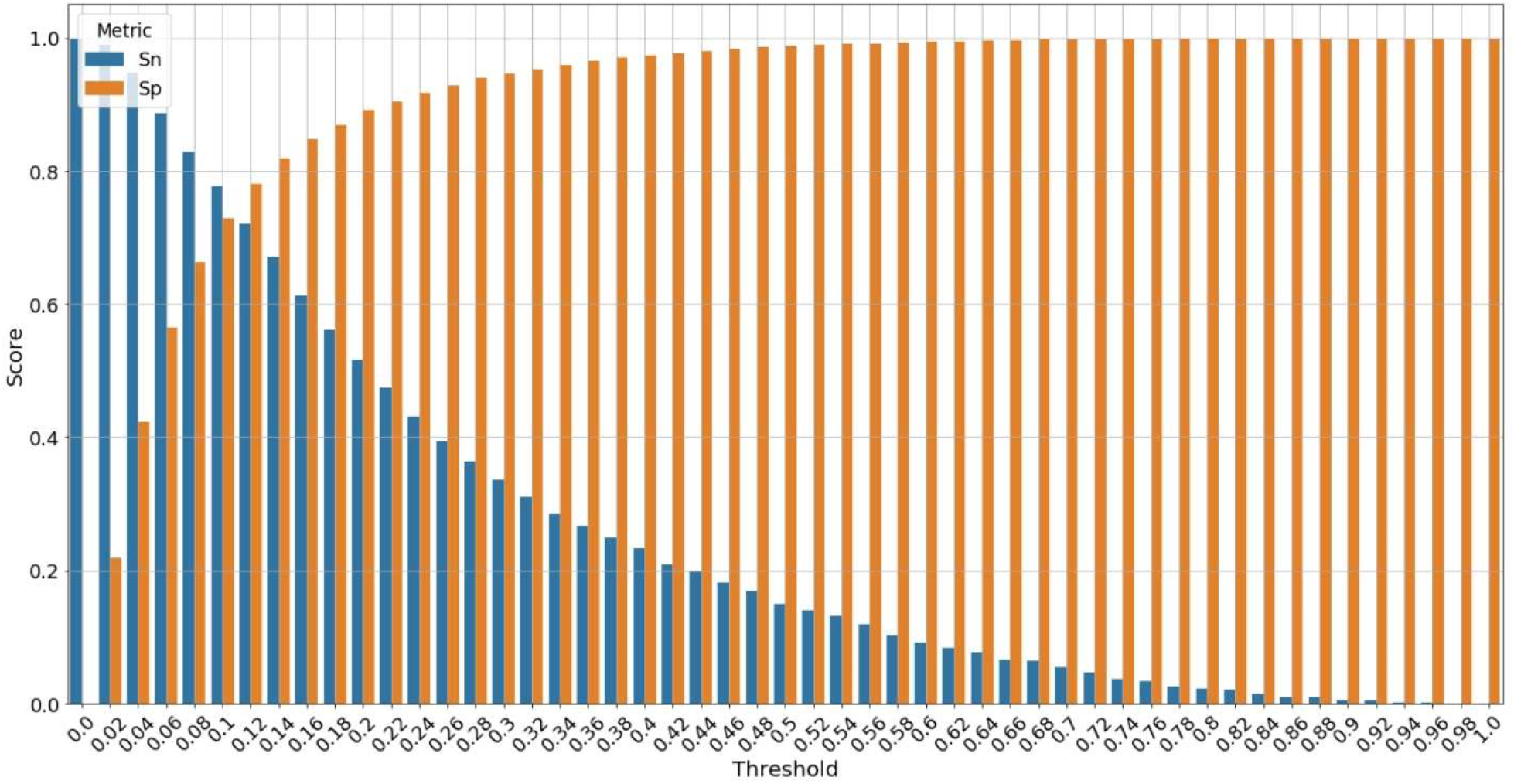
Trade-off between sensitivity and specificity for different choices of the threshold for discretizing the continuous predicted probability into a predicted binary label.

**Fig 6.**
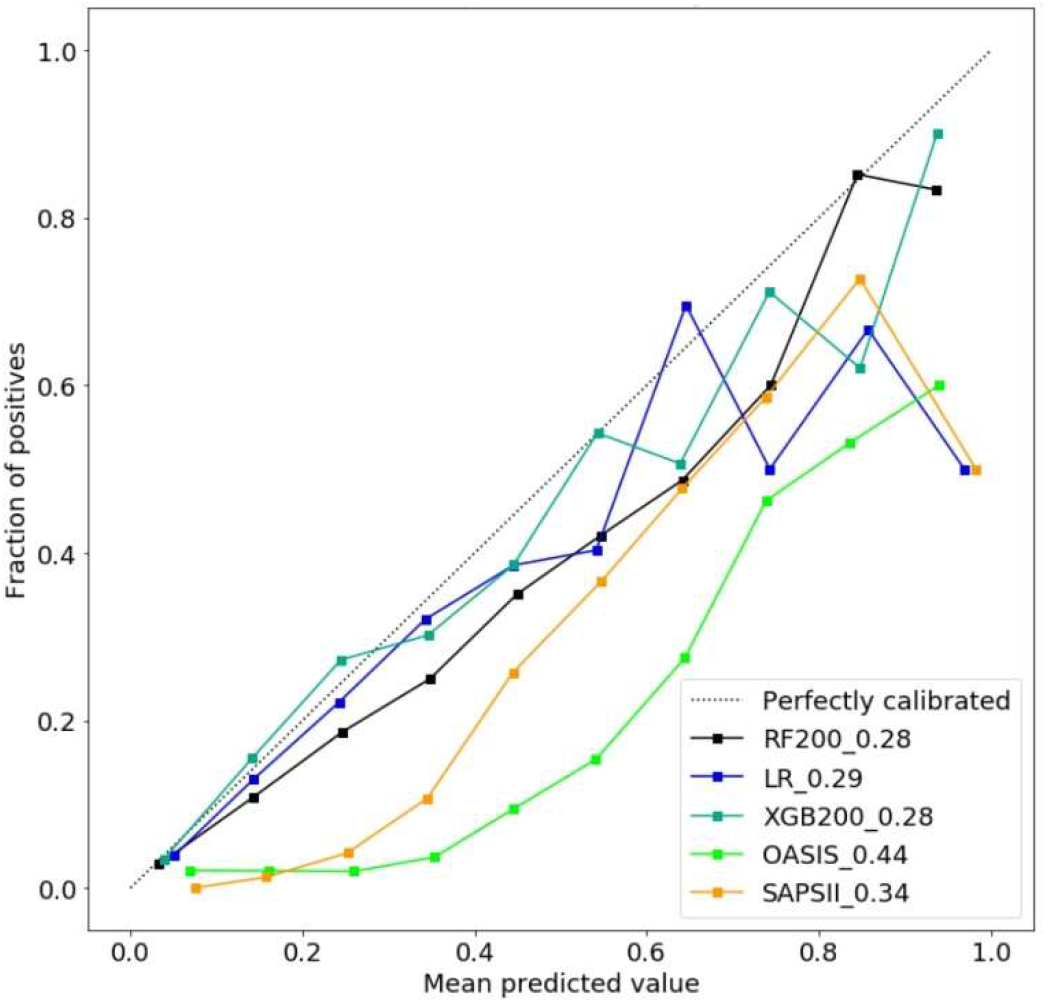
Calibration curves assessing the consistency between the actual risk and predicted risk of different models.

### OASIS+ web application

We deployed the OASIS+ model using the streamlit framework, https://github.com/streamlit/streamlit, and made it publicly available at https://oasis-score.herokuapp.com/. The web app provides an interface for the user to input the measured values for the 10 clinical variables and select the values of the threshold for transforming OASIS+ predicted probabilities of in-hospital mortality into a binary label (e.g., deceased vs. survived). The displayed results include: OASIS severity score and probability; interpretation of OASIS score; OASIS+ predicted probability and predicted in-hospital mortality; OASIS+ model interpretation in terms of feature importance scores quantifying the contribution of the 10 clinical variable to the prediction made by the XGB200 classifier (See Fig. 3); and a projection of the user-specified threshold on the ROC curve of the OASIS+ indicating sensitivity and 1-specificity of OASIS+ model at the user-specified threshold estimated using MIMIC-III test set.

## Discussion

The present study introduced, OASIS+, a novel machine learning-based model for predicting in-hospital mortality using the 10 clinical variables used in the OASIS severity score [14]. Our results suggest that there is room for improving the prognostic accuracy of traditional severity scores by replacing the simple linear additive scoring function with more sophisticated non-linear machine learning models such as RF and XGB. Our results also suggest that the two non-linear supervised learning algorithms considered in our experiments can be directly trained using the observed clinical variables without the need for transforming these measurements into subscores.

Over the past two decades several severity scoring systems were subjected to continuous refinements and improvements. For example, SAPS and APACHE scores have three and four versions, respectively. A common pattern in these severity scoring methods with multiple version is that newer versions often have more variables than those used in preceding versions. For example, SAPS scores versions I-III are based on 14, 17, and 20 variables, respectively. Another example is APACHE scores versions I-IV which use 38, 12, 20, and 145 variables, respectively. While adding more relevant physiologic measurements is likely to improve the predictive performance of a severity scoring methods, it makes implementation for use in real-time challenging. Ideally, refinement and improved performance of existing scoring systems could be achieved without adding more variables. This objective can be reached using more sophisticated machine learning algorithms. To the best of our knowledge, OASIS+ is the first study that demonstrates the promise of focusing on the machine learning component of a severity scoring system to significantly improve its predictive performance. This opens up the possibility for improving existing severity scoring methods by adapting the approach presented in this work.

OASIS+ shares several of the advantages of OASIS when compared to other severity scores. Both OASIS and OASIS+ are based on fewer variables than the vast majority of severity scores. Also, these variables are frequently measured during the ICU stay and do not have high rates of missing values. Another major advantage of the OASIS score is that it can be computed manually without the need for informatics support. However, because it uses an ensemble of complex 200 decision trees, OASIS+ cannot be computed manually. To facilitate easy internet-based access to an OASIS+ calculator for individual ICU patients, we have deployed an OASIS+ model as a freely accessible web app for scientific use. For batch computations on large-scale data, we shared OASIS+ model and supplementary Python scripts on a public source code repository which can be accessed at https://bitbucket.org/i2rlab/oasis/src/master/. Therefore, future studies can easily benchmark the performance of OASIS+ using datasets from other health systems.

Taken the availability of large-scale datasets in healthcare systems together with the recent advances in machine learning research, we argue that population-specific severity prediction models should be preferred over traditional severity scoring methods. We believe this for the following reasons. First, our results showed that MIMIC-III specific machine learning models using only 10 clinical variables outperformed nine commonly used severity scoring methods. Several related studies (e.g., [35-37]) have also shown that machine learning models outperform severity scores on predicting in-hospital mortality. Second, developing health system specific (or local) prediction models enables continuous improvements of the model by including more training data (as more data become available), adding new clinical or laboratory variables to the model, or re-training the model using newly developed machine learning algorithms.

Several machine learning based predictive models are treated as “black-boxes”, which are systems that hide their internal logic to the user [38]. Such models often have better predictive accuracy compared with interpretable models such as linear additive models [39]. However, relying on sophisticated non-linear machine learning models trained using large-scale biomedical datasets raises concerns regarding the ethics and trustability of the utility of black-box decision making systems in ICU settings and healthcare in general [40, 41]. These concerns could be addressed in part by using techniques for post-hoc interpretation and explanation of the predictive model [38]. To shed some light on how the OASIS+ model works, we used the XGB200 inferred feature importance scores to quantify the contribution of each of the 10 clinical measurements to the predictions made by OASIS+. We have also inspected the model via the application of sensitivity analysis to examine the effect of the OASIS+ threshold parameter on the sensitivity and specificity of the model. Ultimately, we believe that machine learning predictive models should be held to the same standard as medications or other therapies given to patients. If using a predictive model improves patient outcomes during a prospective clinical trial (for example where patients are randomized to either having the model results available or not), then the model should be adopted clinically regardless of whether the model is fully transparent or “black-box”. As evidence of this, a class of drugs known as beta blockers have been proven to prolong life when given to certain categories of patients with heart failure, but the underlying physiology of these drugs and exactly how they improve survival remain poorly understood [42-44]. Despite this lack of understanding of their mechanism of action, beta blockers are widely used due to their proven efficacy. We believe that machine learning models should be held to the same standard.

The present study has some limitations. First, this is a retrospective study that used a dataset from a single health system. Second, the data used for training our models were extracted from the same source as the test dataset. Third, the inclusion of severity scores in our experiments was restricted by the availability of PostgreSQL scripts for computing these scores from MIMIC-III in the MIMIC code repository [23]. Our ongoing work aims at evaluating the generalizability of OASIS+ on an independent validation set and including other commonly used severity scores such as APACHE-IV [6] in the analysis.

## Conclusions

We have presented a novel machine learning based severity prediction model, OASIS+. OASIS+ is a variant of the OASIS severity score, where the non-linear transformation of the input clinical variables is omitted, and the simple additive function is substituted with an ensemble of 200 non-linear decision trees. Thus, using a machine learning approach we were able to enhance OASIS+ model performance without the need for introducing additional variables beyond the 10 readily available variables used in the OASIS model. In addition to the original OASIS score, OASIS+ outperformed eight other severity scores in predicting in-hospital mortality. Improved OASIS+ performance came with a trade-off in human readability and cannot be computed manually. To address these limitations, we used feature importance and sensitivity analysis to improve the interpretability of the model and we deployed the model as a publicly available web server to enable access to the model. Moreover, we supported the application of OASIS+ model to large-scale datasets by sharing the learned model and necessary Python scripts through a source code repository. Our future work aims at: improving the explanation of OASIS+ model and its predictions; and evaluating OASIS+ using an independent test set.

## Data Availability

The datasets presented in the current study are available in the MIMIC III database (https://physionet.org/content/mimiciii/1.4/).
Our machine learning model and associated Python scripts are freely available at https://bitbucket.org/i2rlab/oasis/.

## Availability of data and materials

The datasets presented in the current study are available in the MIMIC III database (https://physionet.org/content/mimiciii/1.4/).

Our machine learning model and associated Python scripts are freely available at https://bitbucket.org/i2rlab/oasis/.

## Competing interests

The authors declare that they have no competing interests.

## Funding

YE is supported by a startup funding from Geisinger Health System. The funder had no role in the design of the study, collection, analysis, or interpretation of data or the writing of the manuscript.

## Acknowledgements

We are grateful to the researchers at the MIT Laboratory for Computational Physiology and collaborating research groups for making MIMIC-III data and associated code publicly available to the scientific community.

## Supplementary Information

Additional file 1: Supplementary Tables S1-S4

## Notes

### Competing Interest Statement

The authors have declared no competing interest.

### Author Declarations

The public access to the de-identified MIMIC-III database has been approved by the Beth Israel Deaconess Medical Center (BIDMC) and Massachusetts Institute of Technology (MIT) institutional review boards. All data processing and analyses presented in this study have been conducted in accordance with MIMIC-III guidelines and regulations.

## References

1. Bouch DC, Thompson JP: Severity scoring systems in the critically ill. Continuing education in anaesthesia, critical care & pain 2008, 8(5):181–185.

2. Deliberato R, Ko S, Komorowski M, Armengol dLHM, Frushicheva M, Raffa J, Johnson A, Celi L, Stone D: Severity of Illness Scores May Misclassify Critically Ill Obese Patients. Critical care medicine 2018, 46(3):394.

3. Knaus WA, Wagner DP, Draper EA, Zimmerman JE, Bergner M, Bastos PG, Sirio CA, Murphy DJ, Lotring T, Damiano A: The APACHE III prognostic system: risk prediction of hospital mortality for critically III hospitalized adults. Chest 1991, 100(6):1619–1636.

4. Knaus WA, Zimmerman JE, Wagner DP, Draper EA, Lawrence DE: APACHE-acute physiology and chronic health evaluation: a physiologically based classification system. Critical care medicine 1981, 9(8):591–597.

5. Wagner DP, Draper EA: Acute physiology and chronic health evaluation (APACHE II) and Medicare reimbursement. Health care financing review 1984, 1984 (Suppl):91.

6. Zimmerman JE, Kramer AA, McNair DS, Malila FM: Acute Physiology and Chronic Health Evaluation (APACHE) IV: hospital mortality assessment for today’s critically ill patients. Critical care medicine 2006, 34(5):1297–1310.

7. Le Gall J-R, Loirat P, Alperovitch A, Glaser P, Granthil C, Mathieu D, Mercier P, Thomas R, Villers D: A simplified acute physiology score for ICU patients. Critical care medicine 1984, 12(11):975–977.

8. Le Gall J-R, Lemeshow S, Saulnier F: A new simplified acute physiology score (SAPS II) based on a European/North American multicenter study. Jama 1993, 270(24):2957–2963.

9. Metnitz PG, Moreno RP, Almeida E, Jordan B, Bauer P, Campos RA, Iapichino G, Edbrooke D, Capuzzo M, Le Gall J-R: SAPS 3—From evaluation of the patient to evaluation of the intensive care unit. Part 1: Objectives, methods and cohort description. Intensive care medicine 2005, 31(10):1336–1344.

10. Moreno RP, Metnitz PG, Almeida E, Jordan B, Bauer P, Campos RA, Iapichino G, Edbrooke D, Capuzzo M, Le Gall J-R: SAPS 3—From evaluation of the patient to evaluation of the intensive care unit. Part 2: Development of a prognostic model for hospital mortality at ICU admission. Intensive care medicine 2005, 31(10):1345–1355.

11. Le Gall J, Klar J, Lemeshow S: A new way to assess organ dysfunction in the intensive care unit. ICU scoring Group. Jama 1996, 276(10):802–810.

12. Bone RC, Balk RA, Cerra FB, Dellinger RP, Fein AM, Knaus WA, Schein RM, Sibbald WJ: Definitions for sepsis and organ failure and guidelines for the use of innovative therapies in sepsis. Chest 1992, 101(6):1644–1655.

13. Vincent J-L, Moreno R, Takala J, Willatts S, De Mendonça A, Bruining H, Reinhart C, Suter P, Thijs LG: The SOFA (Sepsis-related Organ Failure Assessment) score to describe organ dysfunction/failure. Intensive care medicine 1996, 22(7):707–710.

14. Johnson AE, Kramer AA, Clifford GD: A new severity of illness scale using a subset of acute physiology and chronic health evaluation data elements shows comparable predictive accuracy. Critical care medicine 2013, 41(7):1711–1718.

15. Le Gall J-R: The use of severity scores in the intensive care unit. Intensive care medicine 2005, 31(12):1618–1623.

16. Pirracchio R: Mortality prediction in the icu based on mimic-ii results from the super icu learner algorithm (sicula) project. In: Secondary Analysis of Electronic Health Records. Springer; 2016: 295–313.

17. Awad A, Bader-El-Den M, McNicholas J, Briggs J: Early hospital mortality prediction of intensive care unit patients using an ensemble learning approach. International journal of medical informatics 2017, 108:185–195.

18. Calvert J, Mao Q, Hoffman JL, Jay M, Desautels T, Mohamadlou H, Chettipally U, Das R: Using electronic health record collected clinical variables to predict medical intensive care unit mortality. Annals of Medicine and Surgery 2016, 11:52–57.

19. Breiman L: Random forests. Machine learning 2001, 45(1):5–32.

20. Chen T, Guestrin C: Xgboost: A scalable tree boosting system. In: Proceedings of the 22nd acm sigkdd international conference on knowledge discovery and data mining: 2016; 2016: 785-794.

21. Johnson AE, Pollard TJ, Shen L, Li-Wei HL, Feng M, Ghassemi M, Moody B, Szolovits P, Celi LA, Mark RG: MIMIC-III, a freely accessible critical care database. Scientific data 2016, 3(1):1–9.

22. Bennett CE, Wright RS, Jentzer J, Gajic O, Murphree DH, Murphy JG, Mankad SV, Wiley BM, Bell MR, Barsness GW: Severity of illness assessment with application of the APACHE IV predicted mortality and outcome trends analysis in an academic cardiac intensive care unit. Journal of critical care 2019, 50:242–246.

23. Johnson AE, Stone DJ, Celi LA, Pollard TJ: The MIMIC Code Repository: enabling reproducibility in critical care research. Journal of the American Medical Informatics Association 2018, 25(1):32–39.

24. Douglas K, Douglas S: PostgreSQL: a comprehensive guide to building, programming, and administering PostgresSQL databases: SAMS publishing; 2003.

25. Le Cessie S, Van Houwelingen JC: Ridge estimators in logistic regression. Journal of the Royal Statistical Society: Series C (Applied Statistics) 1992, 41(1):191–201.

26. Pedregosa F, Varoquaux G, Gramfort A, Michel V, Thirion B, Grisel O, Blondel M, Prettenhofer P, Weiss R, Dubourg V: Scikit-learn: Machine learning in Python. the Journal of machine Learning research 2011, 12:2825–2830.

27. Team RC: R: A language and environment for statistical computing. In.; 2013.

28. Baldi P, Brunak S, Chauvin Y, Andersen CA, Nielsen H: Assessing the accuracy of prediction algorithms for classification: an overview. Bioinformatics 2000, 16(5):412–424.

29. Bradley AP: The use of the area under the ROC curve in the evaluation of machine learning algorithms. Pattern recognition 1997, 30(7):1145–1159.

30. Niculescu-Mizil A, Caruana R: Predicting good probabilities with supervised learning. In: Proceedings of the 22nd international conference on Machine learning: 2005; 2005: 625–632.

31. Walsh CG, Sharman K, Hripcsak G: Beyond discrimination: A comparison of calibration methods and clinical usefulness of predictive models of readmission risk. Journal of biomedical informatics 2017, 76:9–18.

32. Elixhauser A, Steiner C, Harris DR, Coffey RM: Comorbidity measures for use with administrative data. Medical care 1998:8-27.

33. Youden WJ: Index for rating diagnostic tests. Cancer 1950, 3(1):32–35.

34. Cook NR: Statistical evaluation of prognostic versus diagnostic models: beyond the ROC curve. Clinical chemistry 2008, 54(1):17–23.

35. Kong G, Lin K, Hu Y: Using machine learning methods to predict in-hospital mortality of sepsis patients in the ICU. BMC Medical Informatics and Decision Making 2020, 20(1):1–10.

36. Silva I, Moody G, Scott DJ, Celi LA, Mark RG: Predicting in-hospital mortality of icu patients: The physionet/computing in cardiology challenge 2012. In: 2012 Computing in Cardiology: 2012: IEEE; 2012: 245–248.

37. Lin K, Hu Y, Kong G: Predicting in-hospital mortality of patients with acute kidney injury in the ICU using random forest model. International journal of medical informatics 2019, 125:55–61.

38. Guidotti R, Monreale A, Ruggieri S, Turini F, Giannotti F, Pedreschi D: A survey of methods for explaining black box models. ACM computing surveys (CSUR) 2018, 51(5):1–42.

39. Wang J, Fujimaki R, Motohashi Y: Trading interpretability for accuracy: Oblique treed sparse additive models. In: Proceedings of the 21th ACM SIGKDD International Conference on Knowledge Discovery and Data Mining: 2015; 2015: 1245–1254.

40. Vellido A: The importance of interpretability and visualization in machine learning for applications in medicine and health care. Neural Computing and Applications 2019:1–15.

41. Bhatt U, Ravikumar P: Building human-machine trust via interpretability. In: Proceedings of the AAAI Conference on Artificial Intelligence: 2019; 2019: 9919–9920.

42. Cadrin-Tourigny J, Shohoudi A, Roy D, Talajic M, Tadros R, Mondésert B, Dyrda K, Rivard L, Andrade JG, Macle L: Decreased mortality with beta-blockers in patients with heart failure and coexisting atrial fibrillation: an AF-CHF substudy. JACC: Heart Failure 2017, 5(2):99–106.

43. Grandi E, Ripplinger CM: Antiarrhythmic mechanisms of beta blocker therapy. Pharmacological research 2019, 146:104274.

44. Kotecha D, Flather MD, Altman DG, Holmes J, Rosano G, Wikstrand J, Packer M, Coats AJ, Manzano L, Boehm M: Heart rate and rhythm and the benefit of beta-blockers in patients with heart failure. Journal of the American College of Cardiology 2017, 69(24):2885–2896.

